# Systemic corticosteroids show no benefit in severe and critical COVID-19 patients in Wuhan, China: A retrospective cohort study

**DOI:** 10.1101/2020.05.11.20097709

**Authors:** Jianfeng Wu, Jianqiang Huang, Guochao Zhu, Yihao Liu, Han Xiao, Qian Zhou, Xiang Si, Hui Yi, Cuiping Wang, Daya Yang, Shuling Chen, Xin Liu, Zelong Liu, Qiongya Wang, Qingquan Lv, Ying Huang, Yang Yu, Xiangdong Guan, Yanbing Li, Krishnarajah Nirantharakumar, KarKeung Cheng, Sui Peng, Haipeng Xiao

## Abstract

**Background:** Systemic corticosteroids are recommended by some treatment guidelines and used in severe and critical COVID-19 patients, though evidence supporting such use is limited.

**Methods:** From December 26, 2019 to March 15, 2020, 1514 severe and 249 critical hospitalized COVID-19 patients were collected from two medical centers in Wuhan, China. We performed multivariable Cox models, Cox model with time-varying exposure and propensity score analysis (both inverse-probability-of-treatment-weighting (IPTW) and propensity score matching (PSM)) to estimate the association of corticosteroid use with the risk of in-hospital mortality among severe and critical cases.

**Results:** Corticosteroids were administered in 531 (35.1%) severe and 159 (63.9%) critical patients. Compared to no corticosteroid use group, systemic corticosteroid use showed no benefit in reducing in-hospital mortality in both severe cases (HR=1.77, 95% CI: 1.08-2.89, p=0.023), and critical cases (HR=2.07, 95% CI: 1.08-3.98, p=0.028). In the time-varying Cox analysis that with time varying exposure, systemic corticosteroid use still showed no benefit in either population (for severe patients, HR=2.83, 95% CI: 1.72-4.64, p<0.001; for critical patients, HR=3.02, 95% CI: 1.59-5.73, p=0.001). Baseline characteristics were matched after IPTW and PSM analysis. For severe COVID-19 patients at admission, corticosteroid use was not associated with improved outcome in either the IPTW analysis. For critical COVID-19 patients at admission, results were consistent with former analysis that corticosteroid use did not reduce in-hospital mortality.

**Conclusions:** Corticosteroid use showed no benefit in reducing in-hospital mortality for severe or critical cases. The routine use of systemic corticosteroids among severe and critical COVID-19 patients was not recommended.

## Introduction

The current pandemic of coronavirus disease-19 (COVID-19) has become the most severe global health crisis [1]. At present, the cumulative number of confirmed COVID-19 cases worldwide has exceeded 3 million and is still rising rapidly [2]. Although most of COVID-19 patients reportedly had mild symptoms and good prognosis, the mortality of hospitalized severe and critical cases was 18.2% and 49%, respectively [3,4]. Due to the lack of specific therapies for COVID-19, one of the biggest challenges faced by clinicians in all countries is the clinical management of severe and critical cases with the goal of reducing mortality.

Systemic corticosteroids have been studied extensively with variable and inconsistent results in the treatment of acute respiratory distress syndrome (ARDS) caused by viral pneumonia [5-7]. During the outbreaks of severe acute respiratory syndrome (SARS) and Middle East respiratory syndrome (MERS), systemic corticosteroids were used in 79.6% and 48.9% of critical cases, respectively [5,8]. Two studies of patients with SARS and influenza A (H1N1) viral pneumonia showed that the use of systemic corticosteroids was associated with reduced mortality in critical patients [6,8]. However, several other studies of patients with SARS and one study of patients with MERS all indicated that its use could be harmful [5,9-12]. A meta-analysis published in 2019 also showed a significant increase in mortality in influenza pneumonia patients treated with systemic corticosteroids [13]. The COVID-19 treatment guidelines released by the China’s National Health Commission recommended low-dose and short-term use of systemic corticosteroids for patients with rapidly worsening conditions [14-16]. Systemic corticosteroids were used in 44.5% of severe COVID-19 patients in China [17]. However, its routine use is not recommended by the World Health Organization (WHO) clinical management guidelines for patients with severe acute respiratory infection when COVID-19 is suspected [18]. In view of the small sample size and the lack of matched controls in all previous studies, large scale studies are urgently needed to provide evidence of harm or benefit for the use of systemic corticosteroids in severe and critical COVID-19 patients.

In the current study, we analysed the clinical data of 1514 severe and 249 critical COVID-19 cases from two medical centres in Wuhan city and investigated the effects of systemic corticosteroids in severe and critical COVID-19 patients.

## Methods

### Study Population

Consecutive inpatients with laboratory confirmed or clinically diagnosed COVID-19 from Wuhan Hankou Hospital and No. Six Hospital of Wuhan between December 26^th^, 2019 and March 15^th^ 2020 were collected in this study. The final follow up date was March 19^th^, 2020. Patients who met any of the following conditions were exclude from the study: 1. Non severe or critical cases; 2. Not being diagnosed as severe cases within 24 hours since admission; 3. The time of being diagnosed of severe/critical cases were missing. Severe cases were defined as those who required oxygen therapy during hospital stay. Critical cases were those who met any of the following conditions during the whole hospital stay: 1. mechanical ventilation was required; 2. required treatment in intensive care unit (ICU); 3. shock occurred in hospital [16]. This study was approved by the ethics committees of Wuhan Hankou Hospital, No. Six Hospital of Wuhan and the First Affiliated Hospital of Sun Yat-sen University, and the informed consent was waived. The study followed the tenets of the Declaration of Helsinki and is reported as per the Strengthening the Reporting of Observational Studies in Epidemiology (STROBE) guideline.

### Data Collection and Definition

We collected the patients’ clinical data, including demographic information, medical history, laboratory indexes, corticosteroid use and prognosis. The admission laboratory indexes were defined as the first records of laboratory indexes since admission. Corticosteroid use was defined as the use of intravenous systemic corticosteroids, including hydrocortisone, methylprednisolone, and dexamethasone. The dose of corticosteroids was converted to methylprednisolone-equivalent doses (1mg methylprednisolone = 0.1875mg dexamethasone = 5mg hydrocortisone).

### Exposure and Outcome

We evaluated whether using corticosteroids could affect the in-hospital mortality of severe/critical cases. The time to death was defined as the time of being diagnosed of severe/critical cases to the date of death from any cause in hospital.

### Statistical Analysis

Data were expressed as mean and standard deviation (SD) if normally distributed, and median and interquartile range (IQR) with non-normal distribution, and frequency and percentages for categorical variables. We compared baseline characteristics and outcomes of patients who received corticosteroids and those who did not receive any corticosteroid using Students’ t test, Mann-Whitney test for continuous variables, and chi-square test or Fisher exact test for categorical variables. Survival with or without corticosteroid use was analysed using the Kaplan-Meier method and compared by log-rank test. The 28-day in-hospital mortality and its 95% confidence interval was reported for both corticosteroid use and non-corticosteroid use groups. We evaluated the associations between corticosteroid use and prognosis using two approaches. First, multivariable Cox regression models were applied to evaluate the association between corticosteroid use and mortality. Multivariable analysis was adjusted for age, gender, admission laboratory indexes (including lymphocyte, neutrophil granulocyte, platelet, haemoglobin, glucose, C-reaction protein (CRP), lactate dehydrogenase (LDH), creatinine, aspartate aminotransferase (AST), alanine aminotransferase (ALT), blood urea nitrogen (BUN), pulse oxygen saturation (SpO2), hypertension, diabetes, cancer, chronic obstructive pulmonary disease (COPD), chronic kidney disease (CKD) and smoking history. The proportion of missing values of baseline variables was 4.5%, ranging from 0% to 10.6%. The missing data were imputed by multiple imputation for ten times in the multivariable analysis. Second, in Cox regression models, corticosteroid use was accounted as a time-varying exposure to mitigate immortal time bias. In time-varying analysis, data was reconstructed according to the time of corticosteroid use. Therefore, time from a diagnosis of severe/critical case to time of corticosteroid use was categorized as unexposed. The hazards ratios (HRs) and 95% confidence intervals (CIs) of Cox regression models were reported.

We also performed propensity score driven analysis (inverse-probability-of-treatment-weighting (IPTW) and propensity score matching (PSM)) to account for confounding by indication bias in the above mentioned two approaches [19,20]. Multivariable logistic regression was carried out with corticosteroid use as a binary outcome to obtain the predicted probability of corticosteroid use, which was taken as propensity score (PS). All baseline characteristics were used to construct PS for each three parts of patients. In IPTW, for the group with corticosteroids therapy, the weight was equal to 1/PS, while for the group without corticosteroids therapy, the weight was equal to 1/(1-PS). In PSM, calliper value was set as 20% of the standard deviation of propensity scores. We reported the results of multivariable Cox regression, also Cox regression with time-varying exposure before matching and after matching (IPTW and PSM) for all association analysis. Wilcoxon signed-rank test was used for pairwise comparison of mean blood glucose and lymphocytes before, during and after the use of corticosteroids, and method of Bonferroni was used for correction. The distribution of the length of corticosteroid use was plotted by bar chart for both severe and critical patients. We used Stata/MP 14.0 to conduct all data analyses, and statistical significance was set as p<0.05 bilaterally.

## Results

### Baseline characteristics

From Dec 26^th^, 2019 to Mar 15^th^, 2020, a total of 2289 consecutive cases of COVID-19 hospitalized patients were collected from two medical centres in Wuhan, China. After excluding non-severe/critical cases (n=126), cases without the exact severe or critical diagnosis timepoint (n=202) and severe or critical cases not being diagnosed on the day of admission (n=198), a total of 1763 COVID-19 hospitalized severe or critical cases were included. Among them, 85.9% (1514/1763) patients were severe cases at admission, and 14.1% (249/1763) patients were critical cases at admission.

Table 1 showed the baseline characteristics of 1514 severe cases at admission. The median age was 61.0 years (IQR: 51.0, 70.0), and 790 (52.2%) were female. The median in-hospital stay was 12.7 days (IQR: 7.5, 19.8). Among them, 531 (35.1%) patients received systemic corticosteroids with a daily average dose equivalent to 40.0 mg (IQR: 37.3, 57.1) methylprednisolone, and 67.6% (359/531) started corticosteroid use within 24 hours after being diagnosed as severe cases. The median initial time of corticosteroid use since being diagnosed as severe cases was 2.2 (0.1, 41.5) hours, and the duration of corticosteroid use lasted 6.0 (3.0, 10.0) days. When comparing the baseline characteristics between corticosteroid use group and non-corticosteroid use group, there were significant differences in age, gender, several admission laboratory values and smoking (p<0.05).

**Table 1.** Demographics and baseline characteristics of 1514 severe COVID-19 patients at admission.

| **Characteristic** | **Total**<br>**(n=1514)** | **No Corticosteroid**<br>**(n=983)** | **Corticosteroid**<br>**(n=531)** | **P value** |
| --- | --- | --- | --- | --- |
| **Age, y** | 61.0 (51.0, 70.0) | 60.0 (50.0, 69.0) | 63.0 (53.0, 71.0) | 0.001 |
| **Female, n (%)** | 790 (52.2) | 550 (56.0) | 240 (45.2) | <0.001 |
| **Comorbid conditions, n (%)** |  |  |  |  |
| Diabetes | 181 (12.0) | 110 (11.2) | 71 (13.4) | 0.214 |
| Hypertension | 354 (23.4) | 232 (23.6) | 122 (23.0) | 0.799 |
| COPD | 42 (2.8) | 26 (2.6) | 16 (3.0) | 0.743 |
| Cancer | 22 (1.5) | 17 (1.7) | 5 (0.9) | 0.266 |
| CKD | 29 (1.9) | 21 (2.1) | 8 (1.5) | 0.439 |
| Smoking | 185 (12.2) | 107 (10.9) | 78 (14.7) | 0.033 |
| **Admission laboratory values** |  |  |  |  |
| Admission glucose, mmol/L | 6.0 (5.2, 7.5) | 5.8 (5.0, 6.8) | 6.8 (5.7, 8.4) | <0.001 |
| Lymphocyte counts, 10<sup>9</sup>/L | 1.0 (0.7, 1.4) | 1.2 (0.8, 1.5) | 0.7 (0.5, 1.0) | <0.001 |
| Neutrophil counts, 10<sup>9</sup>/L | 3.7 (2.6, 5.2) | 3.6 (2.6, 4.7) | 4.1 (2.7, 6.5) | <0.001 |
| Platelet counts, 10<sup>9</sup>/L | 199.0 (151.0, 251.8) | 211.0 (163.0, 257.0) | 176.0 (138.0, 233.0) | <0.001 |
| Haemoglobin, g/L | 127.0 (116.0, 137.0) | 126.0 (115.0, 136.0) | 129.0 (118.0, 139.0) | 0.001 |
| CRP, mg/L | 24.8 (4.8, 47.8) | 13.8 (2.0, 42.3) | 35.7 (20.9, 64.7) | <0.001 |
| SpO<sub>2</sub>, % | 97.0 (95.0, 99.0) | 98.0 (96.0, 99.0) | 96.0 (92.0, 98.0) | <0.001 |
| Lactate, mmol/L | 1.6 (1.3, 2.1) | 1.6 (1.3, 2.1) | 1.6 (1.3, 2.2) | 0.471 |
| LDH, U/L | 224.6 (179.0, 310.0) | 202.1 (166.0, 266.1) | 285.0 (212.6, 375.6) | <0.001 |
| AST, U/L | 26.0 (18.2, 38.2) | 23.0 (16.9, 34.0) | 31.0 (23.8, 46.0) | <0.001 |
| ALT, U/L | 24.2 (15.9, 37.0) | 23.0 (15.0, 36.0) | 26.0 (18.0, 39.6) | <0.001 |
| D-Dimer, mg/L | 0.4 (0.2, 0.8) | 0.4 (0.2, 0.7) | 0.5 (0.2, 1.1) | <0.001 |

| Characteristic | Total<br>(n=1514) | No Corticosteroid<br>(n=983) | Corticosteroid<br>(n=531) | P value |
| --- | --- | --- | --- | --- |
| Creatinine, $\mu\text{mol/L}$ | 69.2 (57.3, 83.0) | 67.4 (56.1, 80.8) | 72.0 (60.0, 89.0) | <0.001 |
| BUN, mmol/L | 4.4 (3.5, 5.7) | 4.2 (3.4, 5.5) | 4.7 (3.8, 6.3) | <0.001 |
| **Medications** |  |  |  |  |
| The initial time of corticosteroid use since<br>being diagnosed as severe cases, h | - | - | 2.2 (0.1, 41.5) | - |
| $\leq 24$ hours | - | - | 359 (67.6) | - |
| 24-48 hours | - | - | 56 (10.5) | - |
| 48-72 hours | - | - | 38 (7.2) | - |
| >72 hours | - | - | 78 (14.7) | - |
| Accumulative corticosteroid dose, mg* | - | - | 280.0 (140.0, 480.0) | - |
| Duration of corticosteroid use, d | - | - | 6.0 (3.0, 10.0) | - |
| Daily average corticosteroid dose, mg | - | - | 40.0 (37.3, 57.1) | - |
| **Disease severity** |  |  |  |  |
| In-hospital stay, d | 12.7 (7.5, 19.8) | 11.5 (6.9, 17.8) | 15.2 (9.1, 23.8) | <0.001 |
| Progression to critical cases, n (%) | 253 (16.7) | 104 (10.6) | 149 (28.1) | <0.001 |
| Death, n (%) | 109 (7.2) | 26 (2.6) | 83 (15.6) | <0.001 |

\* The dose of corticosteroids was converted to methylprednisolone-equivalent doses.
Abbreviations: COVID-19, coronavirus disease 2019; COPD, chronic obstructive pulmonary disease; CKD, chronic kidney disease; CRP, C-reactive protein; LDH, lactate dehydrogenase; AST, aspartate aminotransferase; ALT, alanine aminotransferase; BUN, blood urea nitrogen.

Table 2 showed the baseline characteristics of 249 critical cases at admission. The median age was 68.0 years (IQR: 58.0, 78.0), and 102 (41.0%) were female. The median in-hospital stay was 13.9 days (IQR: 5.8, 22.6). Among them, 159 (63.9%) patients received systemic corticosteroids with a daily average dose equivalent to 40.0 mg (IQR: 40.0, 60.0) methylprednisolone, and 79.9% (127/249) started corticosteroid use within 24 hours after being diagnosed as critical cases. The median initial time of corticosteroid use since being diagnosed as critical cases was 0.1 (0, 16.1) hours, and the duration of corticosteroid use lasted 5.0 (3.0, 7.0) days. When comparing the baseline characteristics between corticosteroid use group and non-corticosteroid use group, there were significant differences in several admission laboratory values (p<0.05).

**Table 2.** Demographics and baseline characteristics of 249 critical COVID-19 patients at admission.

| **Characteristic** | **Total**<br>**(n=249)** | **No Corticosteroid**<br>**(n=90)** | **Corticosteroid**<br>**(n=159)** | **P value** |
| --- | --- | --- | --- | --- |
| **Age, y** | 68.0 (58.0, 78.0) | 67.0 (54.0, 82.0) | 68.0 (60.0, 75.0) | 0.340 |
| **Female, n (%)** | 102 (41.0) | 37 (41.1) | 65 (40.9) | >0.999 |
| **Comorbid conditions, n (%)** |  |  |  |  |
| Diabetes | 56 (22.5) | 21 (23.3) | 35 (22.0) | 0.875 |
| Hypertension | 107 (43.0) | 39 (43.3) | 68 (42.8) | >0.999 |
| COPD | 24 (9.6) | 12 (13.3) | 12 (7.5) | 0.179 |
| Cancer | 6 (2.4) | 1 (1.1) | 5 (3.1) | 0.423 |
| CKD | 15 (6.0) | 9 (10.0) | 6 (3.8) | 0.056 |
| Smoking | 56 (22.5) | 18 (20.0) | 38 (23.9) | 0.530 |
| **Admission laboratory values** |  |  |  |  |
| Admission glucose, mmol/L | 6.9 (5.7, 8.5) | 6.1 (5.2, 7.7) | 7.3 (6.2, 8.7) | <0.001 |
| Lymphocyte counts, 10<sup>9</sup>/L | 0.7 (0.5, 1.1) | 0.9 (0.6, 1.3) | 0.6 (0.4, 1.0) | <0.001 |
| Neutrophil counts, 10<sup>9</sup>/L | 4.9 (3.5, 8.2) | 4.0 (3.0, 5.8) | 5.9 (3.8, 9.5) | <0.001 |
| Platelet counts, 10<sup>9</sup>/L | 187.0 (141.0, 236.0) | 198.0 (149.0, 246.0) | 182.0 (137.0, 235.0) | 0.215 |
| Haemoglobin, g/L | 126.0 (114.0, 137.0) | 123.7 (109.0, 135.0) | 126.0 (115.3, 138.0) | 0.054 |
| CRP, mg/L | 69.4 (25.7, 140.4) | 47.4 (12.0, 109.0) | 88.1 (34.6, 151.5) | <0.001 |
| SpO<sub>2</sub>, % | 95.0 (90.0, 98.0) | 97.7 (94.9, 99.0) | 94.0 (86.0, 97.2) | <0.001 |
| Lactate, mmol/L | 1.6 (1.3, 2.3) | 1.5 (1.2, 2.0) | 1.7 (1.3, 2.4) | 0.062 |
| LDH, U/L | 331.0 (208.0, 533.2) | 222.8 (162.5, 344.2) | 408.0 (265.0, 596.7) | <0.001 |
| AST, U/L | 33.8 (21.0, 53.0) | 27.1 (19.3, 42.6) | 37.5 (22.3, 57.5) | 0.001 |
| ALT, U/L | 25.7 (16.6, 41.8) | 21.2 (15.5, 35.0) | 27.2 (17.6, 44.0) | 0.045 |
| D-Dimer, mg/L | 0.7 (0.4, 2.1) | 0.5 (0.4, 1.0) | 0.8 (0.4, 3.1) | 0.008 |

| Characteristic | Total<br>(n=249) | No Corticosteroid<br>(n=90) | Corticosteroid<br>(n=159) | P value |
| --- | --- | --- | --- | --- |
| Creatinine, µmol/L | 75.7 (61.0, 104.0) | 75.6 (58.3, 101.8) | 75.8 (61.2, 104.0) | 0.878 |
| BUN, mmol/L | 5.6 (4.2, 8.5) | 5.4 (3.9, 7.8) | 5.6 (4.4, 9.2) | 0.177 |
| **Medications** |  |  |  |  |
| The initial time of corticosteroid use since<br>being diagnosed as critical cases, h | - | - | 0.1 (0.0, 16.1) | - |
| ≤24 hours | - | - | 127 (79.9) | - |
| 24-48 hours | - | - | 7 (4.4) | - |
| 48-72 hours | - | - | 6 (3.8) | - |
| >72 hours | - | - | 19 (11.9) | - |
| Accumulative corticosteroid dose, mg* | - | - | 240.0 (120.0, 360.0) | - |
| Duration of corticosteroid use, d | - | - | 5.0 (3.0, 7.0) | - |
| Daily average corticosteroid dose, mg | - | - | 40.0 (40.0, 60.0) | - |
| **Disease severity** |  |  |  |  |
| In-hospital stay, d | 13.9 (5.8, 22.6) | 15.6 (7.9, 24.5) | 12.9 (5.1, 21.9) | 0.203 |
| Death, n (%) | 84 (33.7) | 14 (15.6) | 70 (44.0) | <0.001 |

\* The dose of corticosteroids was converted to methylprednisolone-equivalent doses.
Abbreviations: COVID-19, coronavirus disease 2019; COPD, chronic obstructive pulmonary disease; CKD, chronic kidney disease; CRP, C-reactive protein; LDH, lactate dehydrogenase; AST, aspartate aminotransferase; ALT, alanine aminotransferase; BUN, blood urea nitrogen.

### Systemic corticosteroid use showed no benefit in reducing in-hospital mortality in severe cases

For all 1514 severe cases, we analysed the factors associated with in-hospital mortality. Kaplan-Meier survival curve showed that in-hospital mortality was significantly higher in the corticosteroid use group than in the no corticosteroid use group (log-rank test p<0.001). The 28-day in-hospital mortality was 20.6% (95% CI: 16.5%-25.6%) in the corticosteroid use group while 3.7% (95% CI: 2.3%-6.0%) for no corticosteroid use. The results of univariate regression model were shown in Supplementary Table 1. In the multivariable Cox model, systemic corticosteroid use was independently associated with in-hospital mortality (HR=1.77, 95% CI: 1.08-2.89, p=0.023) (Table 3). In the multivariable Cox model with time-varying exposure, systemic corticosteroid use was independently associated with in-hospital mortality (HR=2.83, 95% CI: 1.72-4.64, p<0.001). After IPTW and PSM (baseline shown in Supplementary Tables 2 and 3), the results showed tendency towards the association between systemic corticosteroid use and increased in-hospital mortality after both propensity score analysis (HR=1.43, 95% CI: 0.82-2.49, p=0.201 in IPTW; HR=1.55, 95% CI: 0.83-2.87, p=0.166 in PSM). Kaplan-Meier survival curves after IPTW and PSM were shown in Figure 1A and 1B.

**Figure 1.**
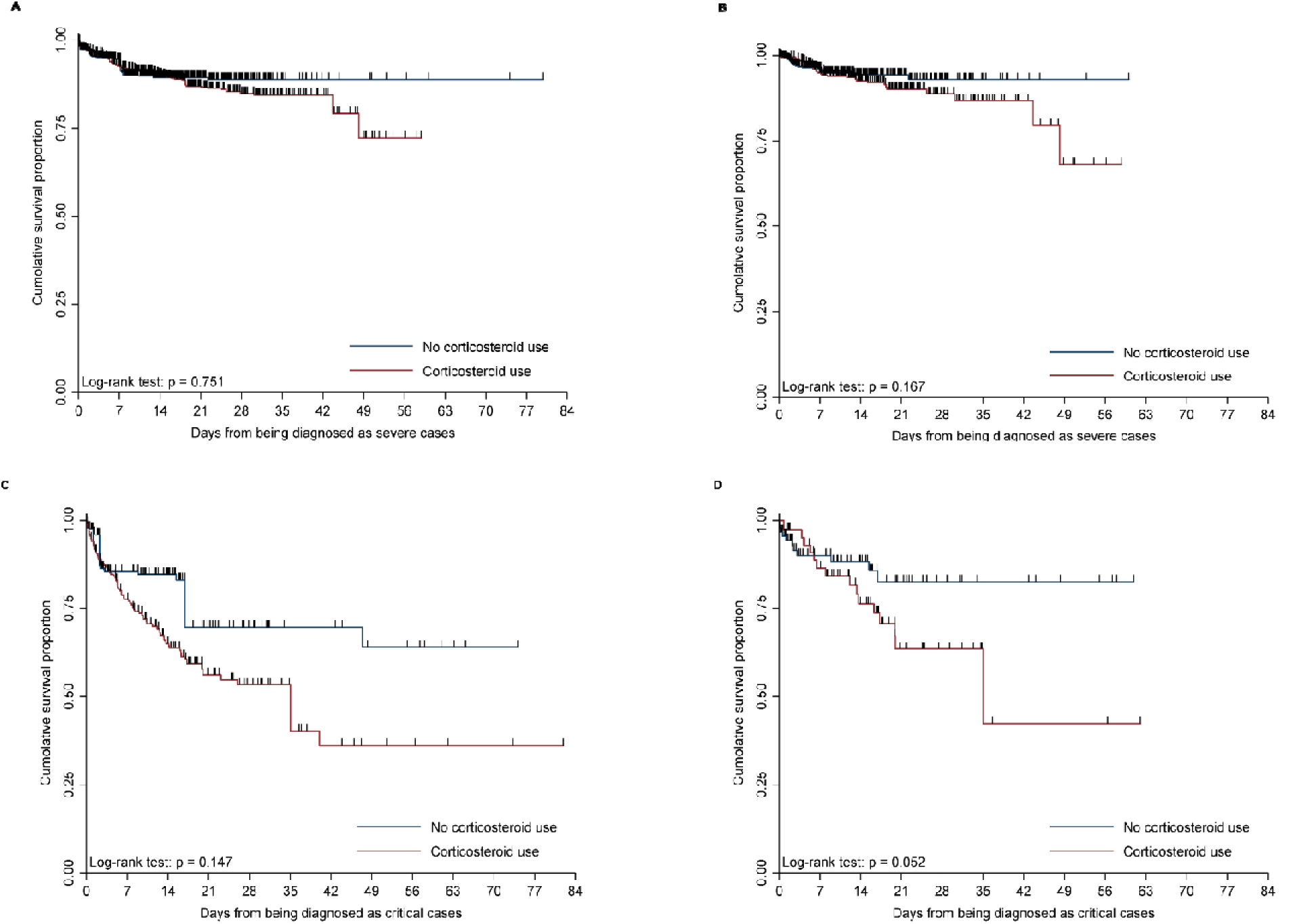
Kaplan-Meier survival curves of the corticosteroid use group and no corticosteroid use group for IPTW and PSM in-hospital mortality in severe cases (A,B), and in critical cases (C,D), respectively. Abbreviations: IPTW, inverse-probability-of-treatment-weighting; PSM, propensity score matching.

**Table 3.**
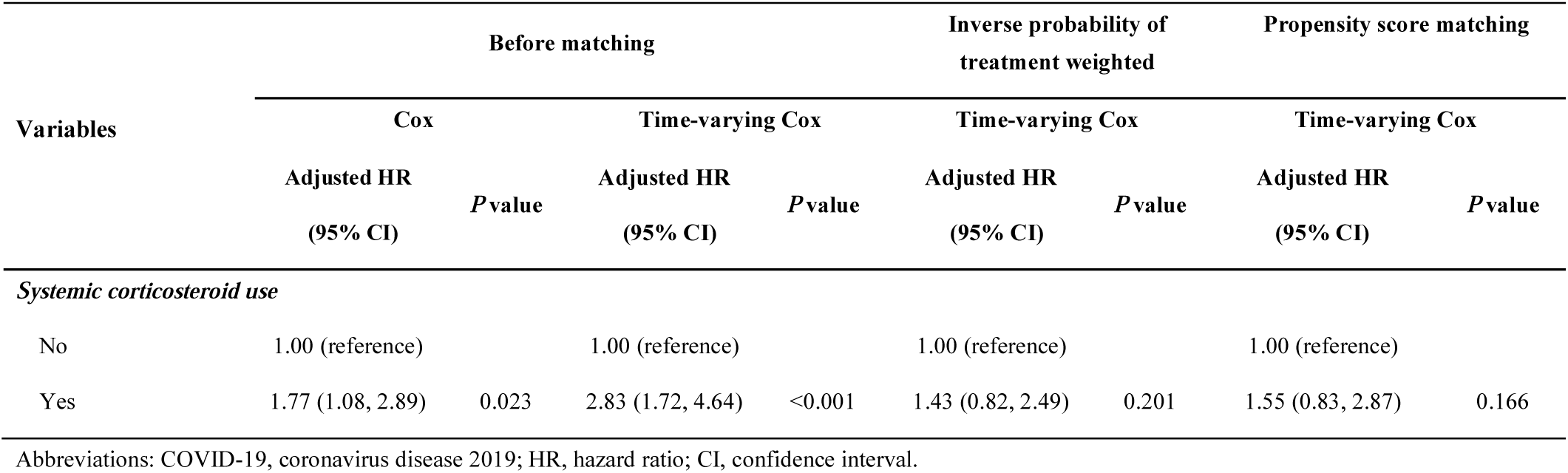
Association of systemic corticosteroid use and in-hospital mortality in severe COVID-19 patients in multivariable Cox regression analysis.

| Variables | Before matching |  |  |  | Inverse probability of treatment weighted |  | Propensity score matching |  |
| --- | --- | --- | --- | --- | --- | --- | --- | --- |
|  | Cox |  | Time-varying Cox |  | Time-varying Cox |  | Time-varying Cox |  |
|  | Adjusted HR<br>(95% CI) | P value | Adjusted HR<br>(95% CI) | P value | Adjusted HR<br>(95% CI) | P value | Adjusted HR<br>(95% CI) | P value |
| *Systemic corticosteroid use* |  |  |  |  |  |  |  |  |
| No | 1.00 (reference) |  | 1.00 (reference) |  | 1.00 (reference) |  | 1.00 (reference) |  |
| Yes | 1.77 (1.08, 2.89) | 0.023 | 2.83 (1.72, 4.64) | <0.001 | 1.43 (0.82, 2.49) | 0.201 | 1.55 (0.83, 2.87) | 0.166 |

Abbreviations: COVID-19, coronavirus disease 2019; HR, hazard ratio; CI, confidence interval.

### Systemic corticosteroid use showed no benefit in reducing in-hospital mortality in critical cases

For all 249 critical cases, we analysed the factors associated with in-hospital mortality. Compared with the no corticosteroid use group, the in-hospital mortality was significantly higher in the corticosteroid use group (log-rank test p<0.001). The 28-day in-hospital mortality was 51.0% (95% CI: 42.2%-60.5%) in the corticosteroid use group and 17.0% (95% CI: 10.0%-28.1 %) in the no corticosteroid use group, respectively. The results of univariate regression model were shown in Supplementary Table 4. In the multivariable Cox model, systemic corticosteroid use was independently associated with increased in-hospital mortality (HR=2.07, 95% CI: 1.08-3.98, p=0.028) (Table 4). In the multivariable Cox model with time-varying exposure, systemic corticosteroid use was also independently associated with increased in-hospital mortality (HR=3.02, 95% CI: 1.59-5.73, p=0.001). After IPTW and PSM (baseline shown in Supplementary Tables 5 and 6), the results supported that systemic corticosteroid use was independently associated with increased in-hospital mortality in critical cases after both propensity score analysis (HR=3.34, 95% CI: 1.84-6.05, p<0.001 in IPTW; HR=2.90, 95% CI: 1.17-7.16, p=0.021 in PSM). Kaplan-Meier survival curves after IPTW and PSM were shown in Figure 1C and 1D.

**Table 4.**
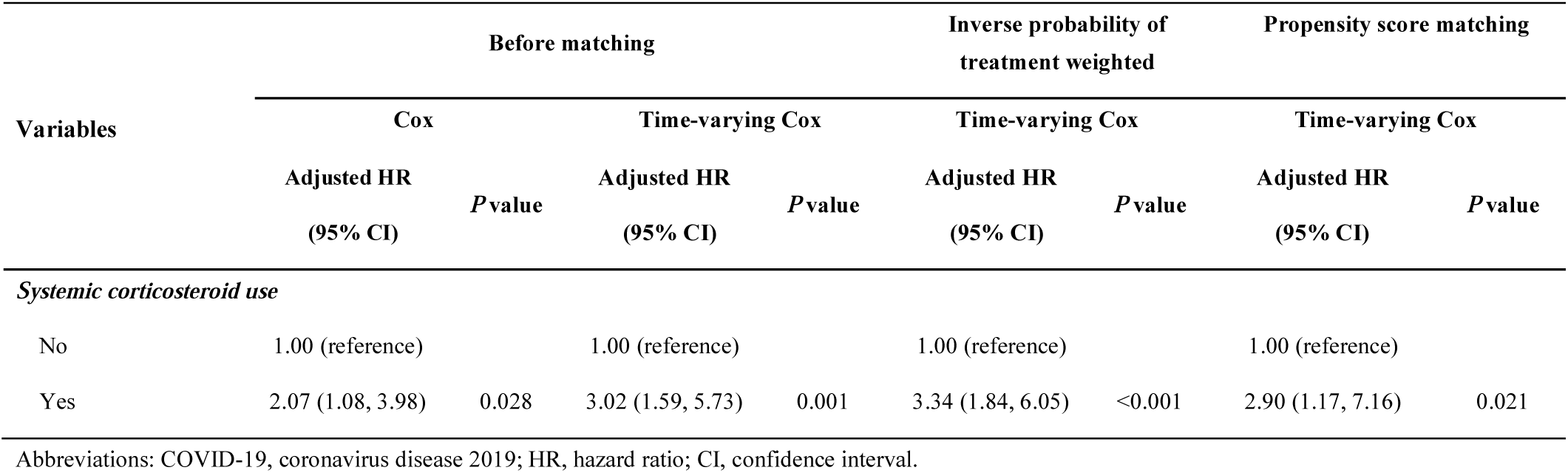
Association of systemic corticosteroid use and in-hospital mortality in critical COVID-19 patients in multivariable Cox regression analysis.

| Variables | Before matching |  |  |  | Inverse probability of treatment weighted |  | Propensity score matching |  |
| --- | --- | --- | --- | --- | --- | --- | --- | --- |
|  | Cox |  | Time-varying Cox |  | Time-varying Cox |  | Time-varying Cox |  |
|  | Adjusted HR<br>(95% CI) | P value | Adjusted HR<br>(95% CI) | P value | Adjusted HR<br>(95% CI) | P value | Adjusted HR<br>(95% CI) | P value |
| *Systemic corticosteroid use* |  |  |  |  |  |  |  |  |
| No | 1.00 (reference) |  | 1.00 (reference) |  | 1.00 (reference) |  | 1.00 (reference) |  |
| Yes | 2.07 (1.08, 3.98) | 0.028 | 3.02 (1.59, 5.73) | 0.001 | 3.34 (1.84, 6.05) | <0.001 | 2.90 (1.17, 7.16) | 0.021 |

Abbreviations: COVID-19, coronavirus disease 2019; HR, hazard ratio; CI, confidence interval.

## Discussion

In the present study, we investigated the effect of systemic corticosteroids on clinical outcomes in 1514 severe and 249 critical COVID-19 cases from Wuhan. Corticosteroids were used in 35.1% and 63.9% in severe and critical cases, respectively. The use of corticosteroids was mainly low-dose and short-term. Corticosteroid use showed no benefit in reducing in-hospital mortality either in severe or in critical COVID-19 cases. The above results were consistent across multivariable Cox regression and Cox model with time-varying exposure; and after propensity score driven analysis (both IPTW and PSM).

Acute respiratory failure was one of the leading causes of death for severe COVID-19 patients. Since there is no specific treatment for coronavirus infection, the current treatment mainly relies on supportive care and oxygen therapy, drawing largely on previous experience in treating SARS and MERS. The use of systemic corticosteroids is one of the most controversial interventions [21]. Two studies on SARS and H1N1 viral pneumonia indicated that use of corticosteroids could reduce mortality in critical patients [6,8]. An observational study by Zhou et al. with 15 COVID-19 patients demonstrated that an improvement in oxygen saturation and arterial partial pressure of oxygen (PaO2)/oxygen fraction (FiO2) was observed 3-5 days after the use of systemic corticosteroids [22], therefore advocating for the use of systemic corticosteroids in patients with severe COVID-19 for a short term. The latest COVID-19 treatment guidelines of the China’s National Health Commission recommended systemic corticosteroid use in severe COVID-19 patients under certain circumstances. However, strong evidence supporting their recommendations is absent. This large observational study found that the use of systemic corticosteroids was not associated with improved outcomes for severe and critical COVID-19 patients.

The rationale for corticosteroid use includes its potential role in suppressing inflammatory storm, reducing inflammatory exudation, and preventing multiple organs injuries in acute respiratory failure. However, its multifaceted negative impacts on prognosis should not be overlooked. Previous studies suggested that the viral load of the novel coronavirus significantly aggravated the severity of the COVID-19 [23]. Systemic corticosteroids may have worsened prognosis by promoting virus replication. Immunity may also be suppressed by the use of systemic corticosteroids [24]. Our results showed that median lymphocyte count remained low during the use of systemic corticosteroids, which might lead to a higher risk of superinfections. Systemic corticosteroids could also induce hyperglycaemia, which was shown to be an independent risk factor for the prognosis of infection and critically ill patients [25-27]. We also found that median blood glucose level was higher during the use of systemic corticosteroids (The change of lymphocyte count and blood glucose level before, during and after corticosteroid use were shown in Supplementary Table 7). In addition, systemic corticosteroids were more likely to cause complications in the elderly – the median age of severe and critical COVID-19 patients being 61.0 and 68.0 years, respectively in this cohort [28,29].

In this study, the baseline characteristics between the corticosteroid use group and the non-corticosteroid use group were unbalanced. In order to reduce the risk of confounding by indication bias, we performed two different methods of propensity score analysis, namely IPTW and PSM. Baseline characteristics were well balanced after matching, but corticosteroid use was not associated with reduced risk of in-hospital mortality. Since patients in the corticosteroid group were unexposed before the initiation of treatment, the use of corticosteroids was also taken as a time-dependent variable to reduce immortality time bias. The results of time-dependent analysis still showed no benefit of the systemic corticosteroids, indicating that our results are robust. Particularly, after matching and using time-dependent analysis, we could observe that the K-M survival curves of corticosteroid use and no corticosteroid use group were close to each other during the first 7 days in both severe and critical cases. The cumulative mortality in first 7 days was similar in two groups, which indicated that the difference in overall mortality was more likely to be caused by the use of systemic corticosteroids, rather than the difference in baseline characteristics.

The major limitation of this study is retrospective nature. Although it was a large scale and the baseline characteristics were balanced between two groups after performing propensity score analysis and time-dependent analysis, our finds should be further validated in randomised controlled trial.

In summary, this large observational study shows that there is no evidence of clinical benefit for the use of corticosteroids in severe and critical COVID-19 patients. We do not recommend its routine use outside a trial setting.

## Data Availability

The datasets generated during and/or analysed during the current study are available from the corresponding author on reasonable request.

## Acknowledgements

We thank all the hospital staff members for their efforts in working in the front line.

## Supplementary material

Supplementary Table 1. Association of variables and in-hospital mortality in severe COVID-19 patients in univariable Cox regression analysis.

Supplementary Table 2. Baseline comparison between corticosteroid use and no corticosteroid use groups after inverse probability of treatment weighted in severe COVID-19 patients.

Supplementary Table 3. Baseline comparison between corticosteroid use and no corticosteroid use groups after propensity score matching in severe COVID-19 patients.

Supplementary Table 4. Association of variables and in-hospital mortality in critical COVID-19 patients in univariable Cox regression analysis.

Supplementary Table 5. Baseline comparison between corticosteroid use and no corticosteroid use groups after inverse probability of treatment weighted in critical COVID-19 patients.

Supplementary Table 6. Baseline comparison between corticosteroid use and no corticosteroid use groups after propensity score matching in critical COVID-19 patients.

Supplementary Table 7. Blood glucose level and lymphocyte counts in severe and critical COVID-19 patients with or without corticosteroid use.

